# Spatiotemporal variation of COVID-19 case time series in the United Kingdom: a dynamic time warping analysis

**DOI:** 10.1101/2025.10.05.25337392

**Authors:** Khai Lin Kong, Freya Shearer, Oliver Eales

## Abstract

**Background:** Analysing the spatiotemporal variation of COVID-19 disease activity can help to identify underlying COVID-19 transmission dynamics and inform the targeting of public health responses. We explored the use of Dynamic Time Warping (DTW), a time series analysis method, to investigate the spatiotemporal variation in COVID-19 disease activity in the United Kingdom (UK) from 1 November 2020 to 19 May 2022.

**Methods:** We performed a DTW analysis on COVID-19 case time series stratified by the 380 Lower Tier Local Authorities (LTLAs) in the UK. First, we performed a hierarchical clustering analysis (HCA) of these time series to investigate their similarity. We then inferred the lead-lag time relationship between time series, both over the entire study period and for three distinct periods of variant emergence (Alpha, Delta and Omicron BA.1).

**Results:** Our clustering analysis found that case time series for LTLAs of Wales, Scotland, Northern Ireland and England were closely related to LTLAs within the same nation. We identified groups of LTLAs in England with highly similar trends in case time series; these groups of LTLAs were geographically clustered despite the methodology not using any geographic information as input. The lead-lag time analysis in England showed that LTLAs in southeast England, the Manchester area, and in London each led the Alpha, Delta, and Omicron BA.1 epidemic waves respectively.

**Conclusion:** We quantified geographic heterogeneity of COVID-19 case time series in the UK using a DTW analysis. The results of our lead-lag time analysis concurred with findings of phylogeographic studies. Further studies to determine DTW’s optimal settings are critical to maximising the potential of DTW in describing the spatiotemporal variation of infectious diseases such as COVID-19.

## Introduction

Infectious disease transmission can be heterogeneous in space and time due to a range of sociocultural, environmental, and pathogen factors(1). International travel, migration and trade can lead to pathogens emerging in different countries at different times. Within countries, distinct small-scale transmission dynamics may occur because of location-specific factors such as demography and mobility networks.

In the United Kingdom (UK), significant variation in SARS-CoV-2 transmission dynamics was observed over the course of the COVID-19 pandemic, including at times, high levels of heterogeneity between geographic regions(2–4). This spatiotemporal variation was likely driven by several factors. For example, the implementation (or relaxation) of non-pharmaceutical interventions (NPIs) decreased (or increased) transmission rates(5), with different stringency and timing between the four nations (England, Wales, Scotland, and Northern Ireland) and between regions of a single nation(6). Other key factors included: the emergence of distinct SARS-CoV-2 variants with different timings of introduction across the UK(7–9), differences in demographics (e.g. population age distribution, population density) between UK regions(4), and the intensity of NPIs(10). To monitor transmission dynamics through time, the UK collected a wide range of surveillance data, including time series of daily SARS-CoV-2 confirmed cases at the lower tier local authority (LTLA) level (380 LTLAs across the UK). Analysing spatiotemporal variation in disease transmission can help to understand underlying drivers of variation and improve the targeting of public health interventions.

Cross-correlation analysis is a common method used to used to analyse epidemic time series data(11–15). It quantifies the similarity between two time series and estimates the lead-lag time relationship (i.e. how much one time series leads or lags another through time) that maximises the correlation between the two series(11,16). However, cross-correlation is limited by its assumption of a constant lead-lag time, which is often not an appropriate assumption for epidemic time series. Dynamic time warping (DTW) is an alternative algorithm that infers the temporal relationship between two time series and allows for the lead-lag relationship to vary through time(17,18). DTW has not been used widely to analyse epidemic time series data, but several examples exist. It has been used to analyse COVID-19 epidemics between regions in the United States (19,20), and between different disease surveillance indicators (e.g. cases, hospitalisation, mobility)(19,21,22). DTW’s ability to analyse time series with a time-varying lead-lag relationship is a useful feature for understanding COVID-19 transmission dynamics where we expect geographic differences in transmission to change through time.

Here we explore the spatiotemporal variation in COVID-19 epidemic activity in the UK from 1 November 2020 to 19 May 2022, using DTW. We quantify the similarity between pairs of COVID-19 case time series from 380 LTLAs across the UK and infer the lead-lag time relationship of each pair. We investigate how the lead-lag patterns across LTLAs changed during periods of novel variant emergence including of Alpha, Delta, and Omicron BA.1 variants.

## Methods

### Data

We retrieved daily COVID-19 case numbers (recorded by the date of specimen collection) from 1 November 2020 to 19 May 2022 for 380 Lower Tier Local Authorities (LTLA) from the United Kingdom Health Security Agency (UKHSA) website(23). We excluded data before 1 November 2020 and after 19 May 2022 (see Supplementary 1). Prior to analysis, each time series, C(t) was transformed to a logarithmic scale, log(C(t) + 1). Each transformed time series was then normalised (mean of zero, standard deviation of one). This ensured that time series from each LTLA — with cases counts varying over multiple orders of magnitude — could be compared. All analyses were performed on the normalised time series in Rstudio (version 4.4.0)(24).

### Dynamic time warping analysis

We calculated the relationship between each pair of time series (accounting for a time-varying lag between them) using a DTW algorithm. A full description of DTW is provided in Supplementary 1. In brief, a DTW algorithm takes two time series and realigns them to minimise the time difference between key features, such as the epidemic peaks (Figure 1). Every time point from one time series is matched to a time point(s) from the other time series following a defined set of rules (see Supplementary 1). Matched time points are said to follow an ‘optimal warping path’, which describes the lead-lag time relationship between a pair of time series. The algorithm does not quantify the uncertainty in this path. The sum of differences between the two time-series along the warping path (i.e. the matched time points) represents the similarity score for that pair.

**Figure 1:**
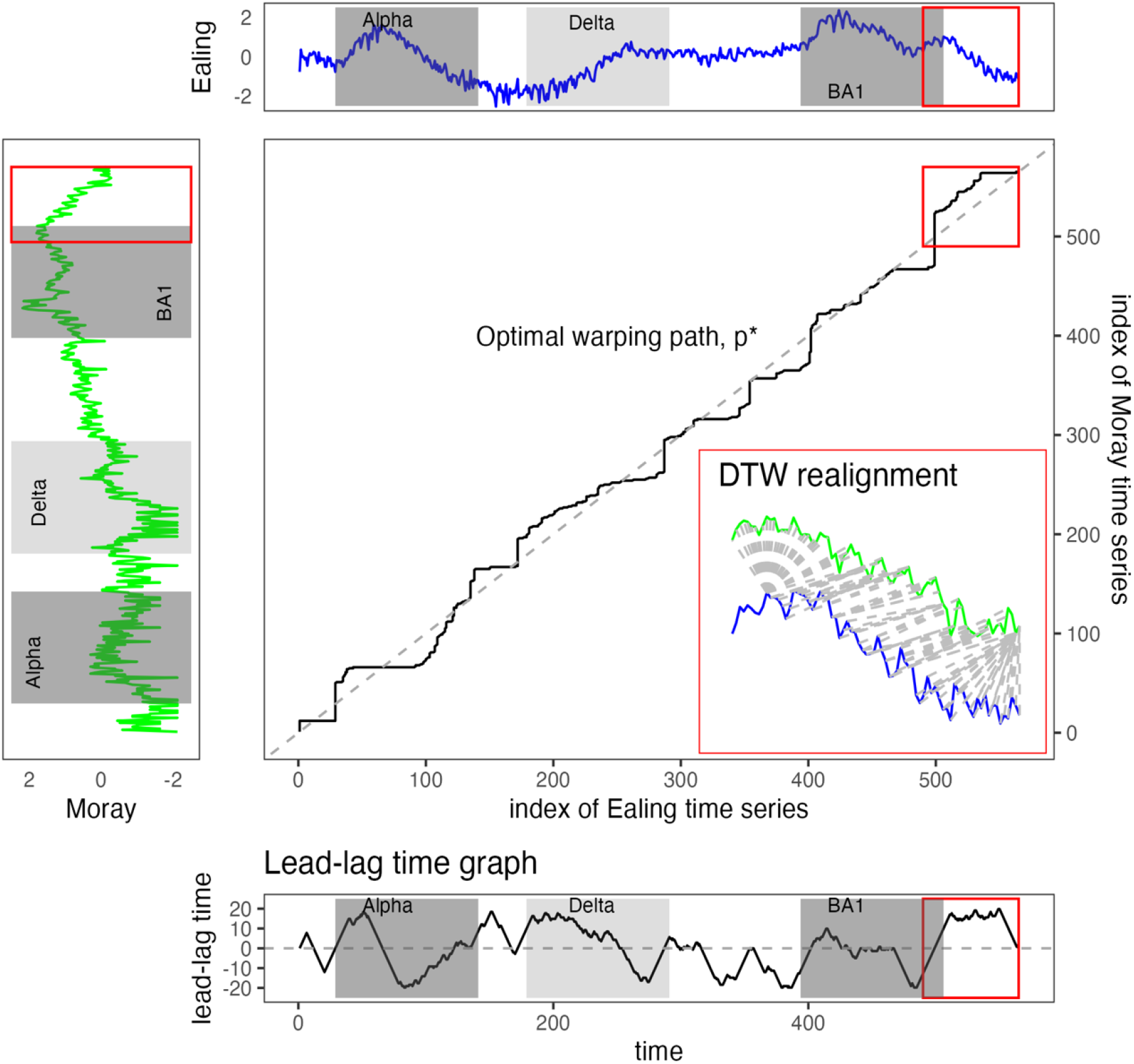
DTW analysis, optimal warping path and lead-lag time analysis for time series from Ealing and Moray. This figure combines the time series for Ealing (**Top**: blue), Moray (**Left**: green), the optimal warping path by DTW (**middle graph**) and a lead-lag time graph between Ealing and Moray (**bottom**). It shows an example of pairwise DTW analysis between the LTLA of Ealing in metropolitan London and the LTLA of Moray in Scotland. The red box titled **‘DTW realignment’** is an example of how DTW algorithm realigns the two sections of Ealing and Moray time series (marked by red box in their respective time series graph). This realignment can also be presented as the ‘optimal warping path’ (middle graph). The coordinates of this optimal warping path consist of the time indices from Ealing and Moray time series matched by the DTW algorithm. (See Supplementary 1 for more on DTW).

We used the DTW algorithm implemented in the dtw R package (version 1.23-1, 2022)(25) and selected the ‘symmetric2’ local step pattern, and Sakoe-Chiba windowing constraints with a window size of 28 days. The Sakoe-Chiba windowing constraints prevent the DTW algorithm from matching time points that are more than 28 days apart since we only intend to consider similarities in temporal trends that are likely due to the same transient epidemic dynamics (i.e. less than 28 days apart). Additionally, we performed all analyses without windowing constraints as a sensitivity analysis (Supplementary 5).

Using similarity measures derived from the DTW analysis of each LTLA time series pair, we performed hierarchical clustering analysis (HCA) across the 380 LTLA time series. For visual interpretation, we displayed the 12 highest level clusters, i.e., LTLAs are represented as 12 clusters, with each cluster comprising LTLAs with case time series more closely related to each other than time series in other clusters. We also obtained the average case time series for each cluster using the DTW Barycenter Averaging (DBA) method available in the dtw_clust package (version 6.0.0, 2024)(26).

We estimated the average lead-lag time relationship across all time series within each nation. Full details are described in the Supplementary 2 and 3. Briefly, for each pair of time series we calculated the lead-lag time, as a function of time, from the optimal warping path (see Figure 1, bottom graph). We calculated the mean lead-lag time for a specific period (see Supplementary 2, Example 2) between each pair of time-series. The relative time lag (RTL) for an LTLA is then the average of the mean lead-lag times between the LTLA and all other LTLAs (see Supplementary 2, Example 3). Time series with a higher value of RTL lead a greater proportion of time series compared to time series with a lower value of RTL. We calculated the RTL across the entire study duration and across three distinct periods reflecting the emergence of novel variants including: (1) Alpha (29 November 2020 to 21 March 2021); (2) Delta (28 April 2021 to 18 August 2021); and (3) Omicron BA.1 (29 November 2021 to 21 March 2022)(Figure 2)(27).

**Figure 2:**
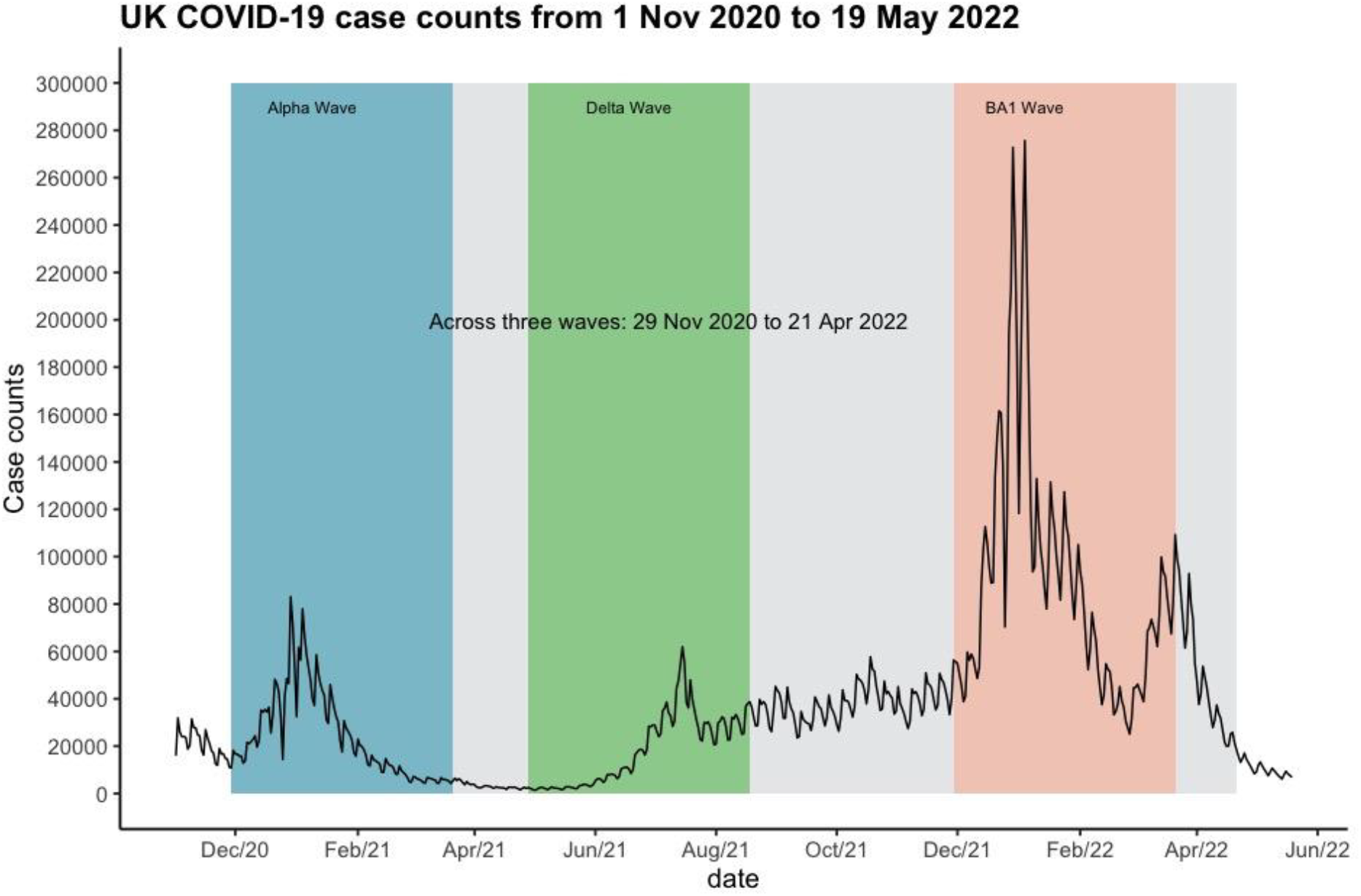
UK COVID-19 case counts. Time series of all COVID-19 cases in the UKHSA from 1 November 2020 to 19 May 2022 (analysis period). The grey (across three waves), blue (Alpha wave), green (Delta wave) and pink (Omicron BA.1 wave) shaded areas represent the four analysis periods used in the lead-lag time analysis.

## Results

### Heterogeneity in case time series between regions

The highest degree of similarity was observed between case time series of LTLAs that were geographically close, despite the methodology not incorporating any measure of geographic proximity (Figure 3). Broad similarities were found between the case time series in Wales (two nested clusters) and between the case time series in Scotland (four nested clusters), which were distinct from the case time series of England and Northern Ireland (six distinct clusters).

**Figure 3:**
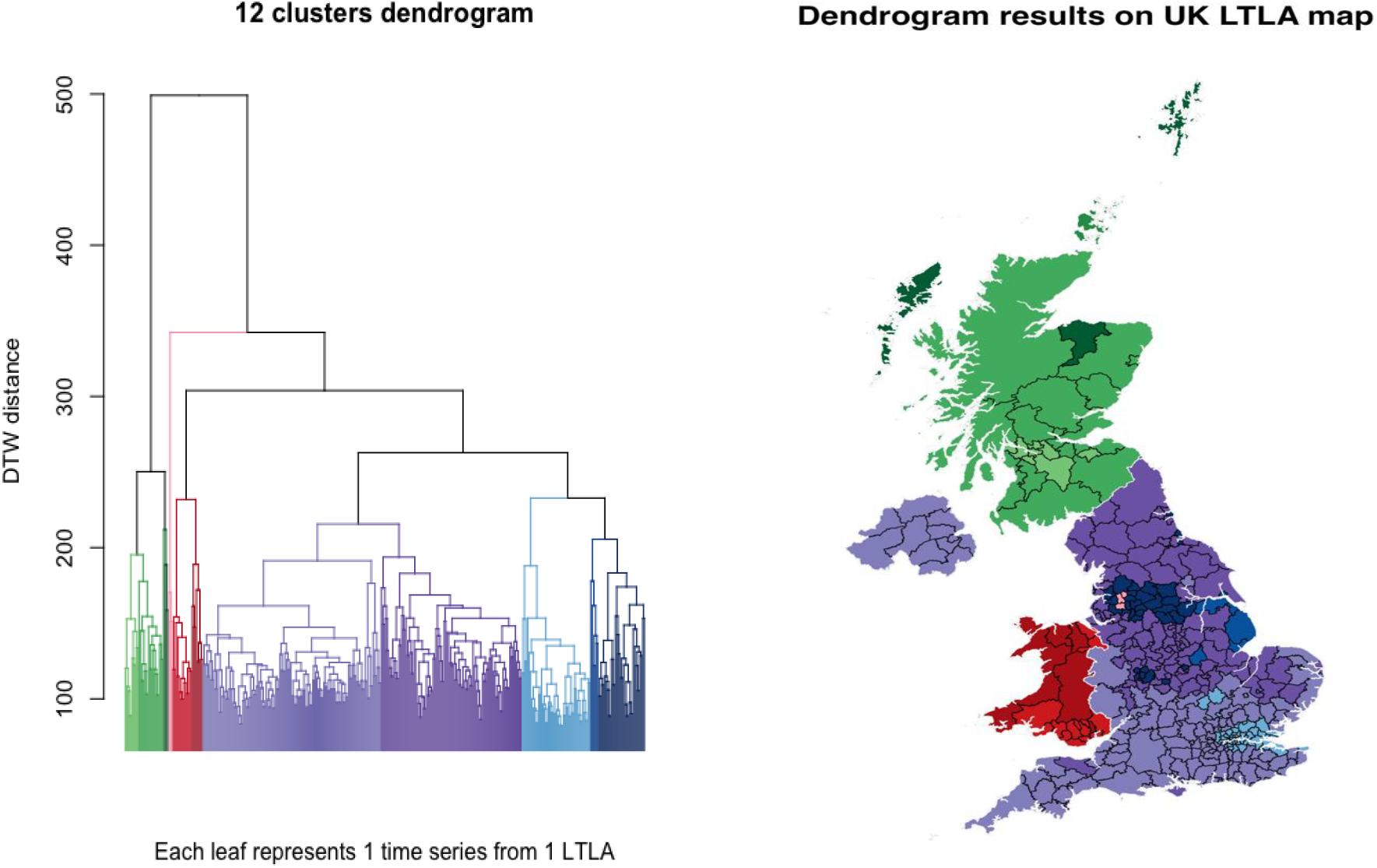
Hierarchical clustering analysis (HCA) **Left:** Dendrogram with each leaf representing a time series from one LTLA. We displayed the HCA output in 12 clusters. The time series within the same cluster are assigned the same colour. **Right:** We displayed the time series on a UK LTLA map based on the location of LTLA from which the time series were collected, and the clusters to which the time series belong. We used the same colour assigned to each time series in the dendrogram to visualise the time series on the map. There are four clusters of time series in Scotland (each coloured in four shades of green). There are two clusters of time series in Wales (coloured in two shades of red). Time series from Northern Ireland are part of the same cluster which also include time series from LTLAs in the South of England (light purple). There are six clusters of time series in England, and they broadly represent time series from (1) LTLAs in and around London (light blue), (2) LTLAs in and around the cities of Manchester, Leeds and Birmingham (dark blue), (3) LTLAs in the South of England (light purple), (4) LTLAs in the North of England (dark purple), (5) LTLAs in the East of England (navy blue), and (6) three LTLAs (Bolton, Blackburn with Darwen and Hyndburn) near Manchester (pink).

Within nations, geographic patterns were observed across LTLAs with similar case time series (i.e. in the same cluster, Figure 3). For example, we found that the time series of South Wales and Central Scotland were similar within each region — representing the more urbanised regions of each nation. Similarly, in England, high levels of similarity were observed between time series from more urbanised areas, with similarities detected between many LTLAs in proximity to London, and between many LTLAs around the cities of Birmingham, Manchester, and Leeds. However, one cluster of LTLAs in England near Manchester (Bolton, Blackburn with Darwen and Hyndburn, Figure 3, pink) was found to be highly dissimilar to all others LTLA in England (i.e. other English LTLAs were more closely related to Welsh LTLAs than this cluster). Across the remaining English LTLAs, a clear North South pattern was observed, with case time series of LTLAs in the South of England and North of England forming two distinct clusters. Case time series in Northern Ireland were closely related to those in England and were most similar to case time series in the South of England.

The average (transformed) case time series for each of the 12 clusters were broadly similar, with some notable differences (Figure 4). For example, from November 2020 until June 2021, the average time series for North Wales was often flat (likely reflecting sustained periods of zero cases), whereas clear temporal dynamics were observed over this period for South Wales. Additionally, for the English cluster that was highly dissimilar to the rest of England (Bolton, Blackburn with Darwen and Hyndburn), a sharp increase was observed in the average case time series from May 2021 to June 2021 during the emergence of the Delta variant. The sharpness of this increase was not observed in the average time series for any of the other clusters.

**Figure 4:**
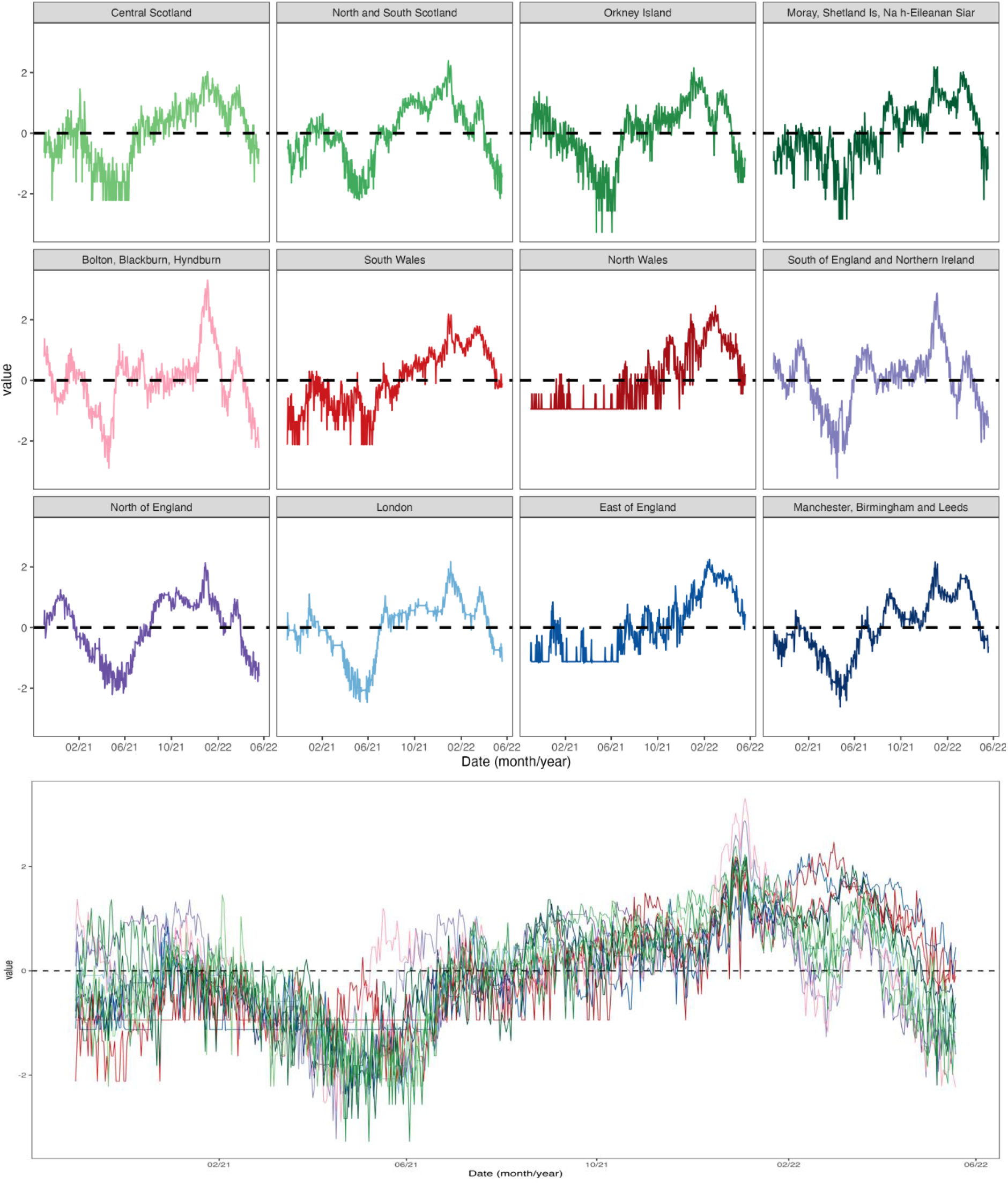
Day Barycenter Averaging (DBA) of time series for each cluster. **Top:** The average time series for each cluster. We used the same colour as the dendrogram in Figure 3 to represent each cluster in Figure 4. We named each cluster based on where the LTLAs of the time series in each cluster are broadly located. The sequence of each plot (left to right and top to bottom) corresponds to the sequence of clusters in the dendrogram (left to right, Figure 3). **Bottom:** The average time series of each cluster plotted in the same graph.

When the DTW analysis was performed without windowing constraints (i.e. time series features greater than 28 days apart can be matched), minor differences were found in the hierarchy of clustering. For example, a greater division was observed between urban and rural areas (Figure S9), with similarities detected between case time series in South Wales and London, and between urbanised areas in the Midlands and North of England (although rural areas also appeared in this cluster). As in the main analysis, case time series in Scotland were found to be closely related. However, in contrast with the main analysis, similarities were detected between the Scottish time series and a few LTLAs in the South East of England. Without windowing constraints, features of two time series that are far apart in time, potentially with very different policy and epidemiological situations, can be matched, which might explain the change in the hierarchy of clustering when this DTW setting is relaxed.

### Lead-lag case dynamics during variant emergence

Our analysis showed that certain geographic regions tended to lead overall case dynamics, but these patterns differed during periods of variant emergence. In England, over the entire course of the study period, case dynamics were broadly led by: LTLAs in London; LTLAs in proximity to London in the South East; and LTLAs in proximity to Manchester in the North West (Figure 5A). However, the degree to which any LTLA led other LTLAs over the entire period was minimal. In contrast, within each period of variant emergence, we found a higher degree of variation in lead times. Specifically, during the emergence of the Alpha variant, case time series of LTLAs in London and the South East were leading other LTLAs (Figure 5B). During the emergence of the Delta variant, case time series of LTLAs in the North West — particularly LTLAs in proximity to Manchester — were leading (Figure 5C). Finally, during the emergence of the Omicron variant, case time series of LTLAs in London were leading, but the lead times were smaller relative to the other periods of variant emergence (Figure 5D). Geographic patterns in lead-lag case dynamics were less clear for Scotland, Wales, and Northern Ireland (Figure S8). These three nations have far fewer LTLAs than England, and so the RTL — which is the average of pairwise comparisons between an LTLA and all other LTLAs — will exhibit greater variation. Our findings were broadly consistent when the DTW analysis was performed without windowing constraints (Figures S10 and S11).

**Figure 5:**
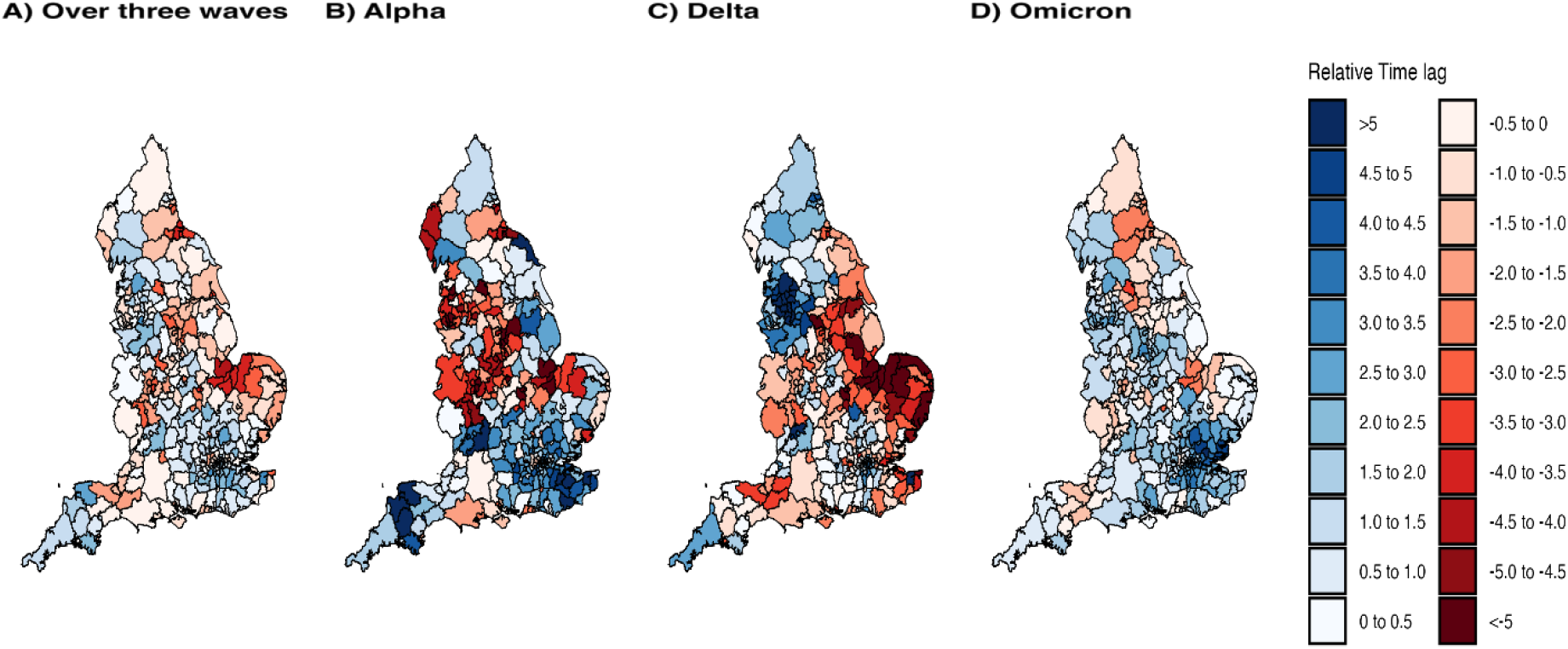
Lead-lag time relationship analysis in England across four periods. **Figure 5A** (first on the left): Analysis period: 29 November 2020 to 21 April 2022 (including the three separate periods when the Alpha, Delta and Omicron BA1 variants each emerged and became the dominant COVID-19 lineage). **Figure 5B** (second from the left): Analysis period: 29 November 2020 to 21 March 2021 (when the Alpha variant was emerging and became the dominant lineage). **Figure 5C** (third from the left): Analysis period 28 April 2021 to 18 August 2021 (when the Delta variant was emerging and became the dominant COVID-19 lineage). **Figure 5D** (fourth from the left): Analysis period 29 November 2021 to 21 March 2022 (when the Omicron BA1 variant was emerging and became the dominant COVID-19 lineage).

## Discussion

We applied a DTW approach to investigate spatiotemporal heterogeneity of COVID-19 case dynamics in the UK from 1 Nov 2020 to 19 May 2022. We found higher degrees of similarity between case time series of LTLAs that were geographically closer. This was despite the DTW approach not using any data on the geographic distance between LTLAs.

Differences in the timing and stringency of public health interventions between the four nations(6) likely contributed to the geographic patterns in case dynamics observed in our analysis. For example, ‘lockdowns’ were implemented for several weeks in late 2020 in England (5 November –2 December 2020)(28), Wales (23 October – 8 November 2020)(29), and Northern Ireland (27 November –11 December 2020)(30), but not in Scotland(31). Similar restrictions for all LTLAs within a nation likely contributed to similar transmission dynamics and, thus, similar case dynamics. Our study found that case time series for LTLAs of Wales, Scotland, Northern Ireland and England were closely related to LTLAs within the same nation. Case time series for LTLAs of Wales, Scotland, and England were distinct from each other. Notably, we did not identify a major difference between case time series of LTLAs in Northern Ireland and those in England. Case time series in Northern Ireland were most similar to those in the South of England, suggesting high levels of similarity in transmission dynamics of SARS-CoV-2 between these two regions. Population demography may also have contributed to geographic patterns in case dynamics. We identified (within-nation) similarities: between LTLAs in London; between LTLAs in Manchester, Birmingham and Leeds; and between LTLAs in South Wales. The higher population density of these areas may have resulted in distinct transmission dynamics relative to areas (in the same nation) with lower population densities(32).

Our analysis detected distinct patterns in case dynamics during the emergence and spread of the Alpha, Delta, and Omicron BA.1 SARS-CoV-2 variants of concern. Bolton and Blackburn with Darwen were two of the first LTLAs where the Delta variant was detected in the UK(33). Our study identified high levels of similarity between the case time series of Blackburn with Darwen, Bolton, and Hyndburn (neighbouring Blackburn with Darwen), and they were distinct from the time series of other LTLAs in England. We defined the relative time lag (RTL) metric to compare the lead-lag time relationship between each time series and all other time series during the emergence of each variant. We found that areas in the southeast of England, and areas in the Greater Manchester region led case dynamics during the emergence of the Alpha and Delta variants respectively. This was in line with phylogeographic studies: the Alpha variant was first detected in southeast England and subsequently spread northward to the rest of the UK(7); and Greater Manchester exported three times more Delta cases to other parts of the UK than Greater London when the Delta variant was emerging(8). During the emergence of the Omicron variant, we estimated that there was less difference in the lead-lag relationship between all LTLAs. The Omicron BA.1 variant was more transmissible than previous variants(34,35) and imported into the UK at a time when social restrictions had been relaxed with high levels of human mobility(9). These two factors likely resulted in the rapid spread of BA.1 infections across the UK and more homogenous case dynamics between regions, as reflected in our lead-lag relationship analysis.

Our findings demonstrate that DTW is a useful tool for quantifying heterogeneity in epidemic time series between geographic regions. By quantifying which regions exhibit similar case dynamics, spatial targeting of public health interventions may be improved, and once those interventions are implemented, DTW analysis may assist in identifying variation in their effectiveness between regions. Additionally, we computed a new metric—the relative time-lag (RTL)—and used it to identify regions with generally leading (or lagging) case dynamics. This type of analysis potentially has implications for real-time epidemic analysis and forecasting. By identifying regions that tend to “lead” other regions in real-time, this may enable improved predictions of epidemic trends in “lagging” regions. Future work could investigate integrating DTW approaches into infectious disease forecasting models.

Dynamic time warping is a relatively new approach for analysing epidemic time series, and as such limited guidelines exist on the appropriate DTW settings to apply. For example, we found that the outcomes of our analysis were sensitive to the selected window size and type. Additionally, there are no standards around transformations that should be applied to epidemic time series before DTW analysis is performed. Previous studies have used varying approaches, including normalising case counts with a z-score(21), normalising case counts with a min-max approach(19), or smoothing the daily case counts with specific variational techniques(22). In contrast to these studies, we used the (normalised) logarithmic value of the daily case counts. This prevents periods of high daily case counts that vary over orders of magnitude from dominating DTW’s realignment process. Normalising the time series also maximises their comparability and prevents DTW from only detecting similarities due to similar *absolute* case numbers as opposed to similar *trends* in case numbers. In the future, analysis of epidemic time series using DTW could consider comparing the growth rates of case time series instead (calculated using statistical models that account for noise in the case time series). The growth rate directly measures the rate at which an epidemic grows or declines, which will more accurately reflect transmission dynamics compared to the logarithm of case counts. Simulation studies could be used to test and compare DTW settings and data transformation options to develop more robust guidelines for the analysis of epidemic time series using DTW.

Our study highlights the importance of collecting high-resolution spatiotemporal surveillance data, ideally with standardised recording and reporting across jurisdictions. Without the availability of daily LTLA-level COVID-19 data, analyses that aim to quantify spatiotemporal patterns in transmission dynamics, such as ours, and ultimately identify underlying drivers of those dynamics, would not be possible. Having insight into the drivers of local transmission dynamics can assist in the spatial targeting and evaluation of public health interventions(36). Subnational data on COVID-19 disease activity are unavailable in some countries(37) and even when available, inconsistent collection of data over time or between regions may complicate spatiotemporal analysis(38). Improved standardisation of surveillance and collection of geographically resolved data have the potential to enhance both national and subnational epidemic analysis, including through methods such as DTW, assisting in the targeting of infectious disease control measures.

## Data Availability

The data that support the findings of this study are available in the United Kingdom Health Security Agency (UKHSA) COVID-19 Archive at https://ukhsa-dashboard.data.gov.uk/covid-19-archive-data-download and Office for National Statistics Open Geography Portal at https://geoportal.statistics.gov.uk/datasets/ons::nuts-level-1-january-2018-boundaries-uk-bfc-2/about.

https://geoportal.statistics.gov.uk/datasets/ons::nuts-level-1-january-2018-boundaries-uk-bfc-2/about.

