## Supplementary 1, 2, 3, 4, and 5 for "Spatiotemporal variation of COVID-19 case time series in the United Kingdom: a dynamic time warping analysis"

### Supplementary 1 (Methods: Data collection, preparation and Dynamic Time Warping)

#### Data collection and preparation

We downloaded daily COVID-19 positive case numbers (recorded by the day of specimen collection from 1 June 2020 to 13 December 2023 for 380 Lower Tier Local Authorities (LTLAs) from the United Kingdom Health Security Agency (UKHSA) website(1). LTLAs are local government authorities that provide a range of local services, equivalent to a Local Government Area (LGA) in Australia. We also downloaded the LTLAs' geospatial data from the Office of National Statistics (ONS) website for mapping purposes(2). The UKHSA COVID-19 database combined the COVID-19 case counts for the following LTLAs: (1) Hackney and City of London; and (2) Cornwall and Isles of Scilly. Consequently, we only used the Hackney geospatial data to map the combined COVID-19 data from Hackney and the City of London, and the Cornwall geospatial data to map the combined COVID-19 data from Cornwall and Isles of Scilly.

We downloaded the COVID-19 data in three separate comma-separated value (CSV) files. We combined these CSV files into a dataset comprising columns of dates and case counts for each LTLA. We excluded the data before 1 November 2020 as the case count for October 2020 was not available for all LTLAs. We also excluded data after 19 May 2022 due to incomplete data for LTLAs in Northern Ireland after 19 May 2022.

Each time series, $\mathrm{C(t)}$ was transformed to a logarithmic scale, $\log(\mathrm{C(t)}+1)$. Each transformed time series was then normalised (mean of zero, standard deviation of one). This ensured that all time series — with cases counts varying over multiple orders of magnitude — could be compared.

#### Dynamic Time Warping (DTW)

DTW algorithm takes two time series and realigns them to minimise the difference between them(3,4). Every time point from one time series is matched to time point(s) from the other time series following a defined set of rules. The time points that are matched by DTW algorithm describe the 'optimal warping path', $(\mathrm{p}^*)$, which minimises the differences between two time series. These minimal distance is also referred to as DTW distance, which is also the minimum cumulative costs, or the minimum sum of differences between the two time series along a warping path(3,4). So, for time series A and B, their DTW distance is:

$\mathrm{DTW(A,B)} = \mathrm{c}_{\mathrm{p}*}\mathrm{(A,B)} = \min\{\mathrm{c_p(A,B)}\}$, where:

1. A and B are time series represented by sequences of values (see Figure S1 and Table S1):
    a. $\mathrm{A} = \{\mathrm{a_1, a_{2,} \ldots a_9}\}$ where $\mathrm{N} = 9$, and i represents the $\mathrm{i^{th}}$ value of A
    b. $\mathrm{B} = \{\mathrm{b_1, b_{2,} \ldots b_9}\}$ where $\mathrm{M} = 9$, and j represents the $\mathrm{j^{th}}$ value of B

2. p represents a warping path. It consists of a sequence of points, $p_l$, with coordinates $(p_i, p_j)$. The $i^{th}$ and $j^{th}$ time indices of X and Y makes up the coordinates $(p_i, p_j)$. We can also represent the optimal warping path as:
   $p^* = (p_1, p_2, \ldots, p_K)$ with $p_l = (p_i, p_j) \in [1:N] \times [1:M]$ for $l \in [1:K]$. See Figure S2.
3. $c_{p*}(X, Y)$ is the distance, also known as the cumulative cost, of the ideal or optimal warping path, $p^*$
4. $c_p(X, Y)$ is the cumulative cost of any warping path

| Time | 1 | 2 | 3 | 4 | 5 | 6 | 7 | 8 | 9 |
|---|---|---|---|---|---|---|---|---|---|
| A | 7 | 9 | 6 | 9 | 12 | 6 | 4 | 6 | 8 |
| B | 5 | 6 | 4 | 3 | 9 | 5 | 6 | 8 | 9 |

**Table S1 Value for time series A and B**

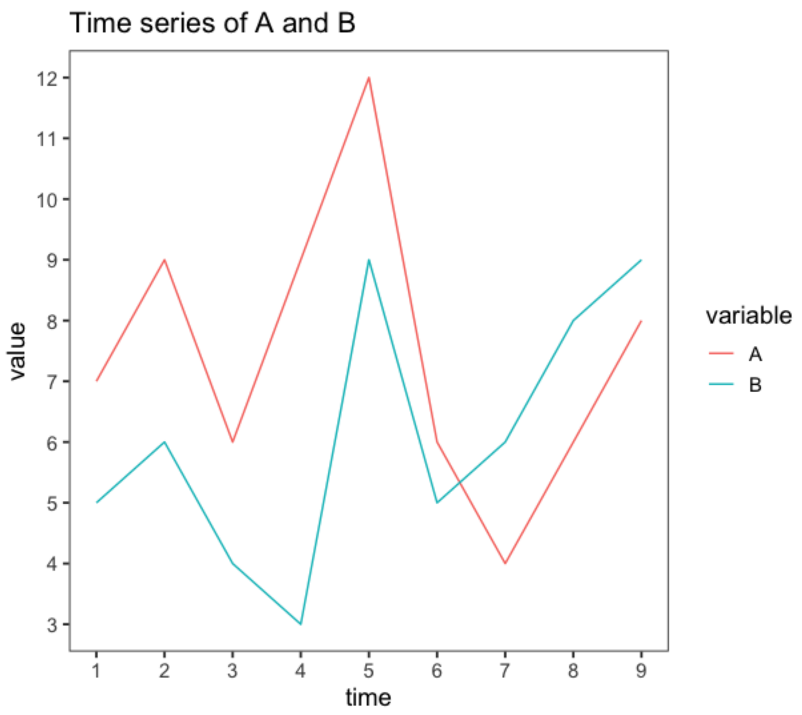


**Figure S1: Time series of A and B**

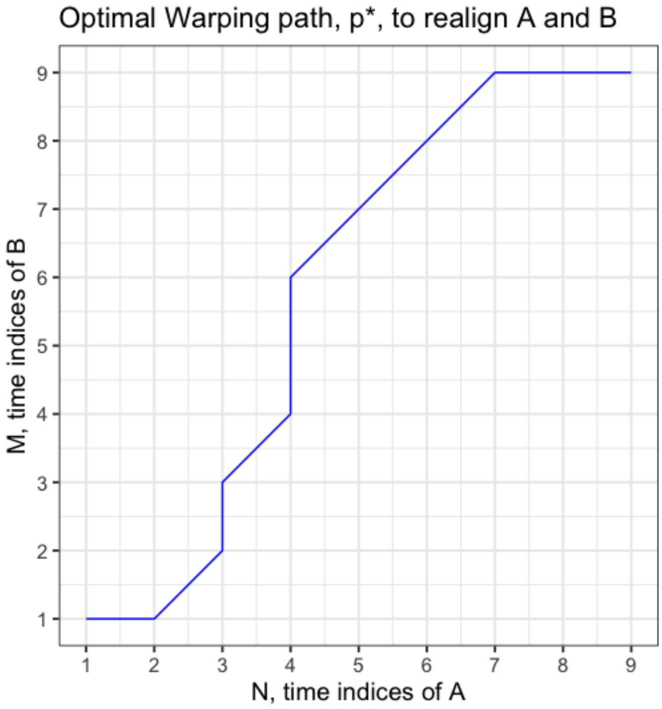


**Figure S2: The optimal warping path**
The blue line shows the optimal warping path to realign A and B. This path consists of a sequence of points. The coordinates of these points are made up of the i$^{th}$ and j$^{th}$ time indices of X and Y matched by the DTW algorithm (where $DTW(A,B) = c_{p*}(A,B) = min\{c_p(A,B)\}$).

#### Step patterns

Step patterns list conditions that determine how the DTW algorithm selects an optimal warping path(3,5). Specifically, step patterns determine how each element of a cumulative cost matrix ($\mathbf{D}^{\mathbf{N}\times\mathbf{M}}$) are calculated. See Figure S5 and Example 1 for an example of cumulative cost matrix and how each element are calculated.

There are several options of step patterns that we can prespecify (e.g. symmetric1, symmetric2, or asymmetric). In this project, we used the ‘symmetric2’ step pattern.  Unlike asymmetric or symmetric1 (a quasi-symmetric step pattern), symmetric2 is symmetrical and does not favour any direction of the warping path(6). See step 3 and 5 in Example 1 on how symmetric2 influences the DTW output of warping path and DTW distance.

#### Windowing constraints

Windowing constraints restrict the selection of warping path to certain parts of the cumulative cost matrix(3). This restriction avoids pathological mappings of two time series when one small section of a time series is matched to a large section of another time series(7). Sakoe-Chiba Band and Itakura Parallelogram are two commonly used windowing constraints. Sakoe-Chiba Band limits the selection of warping path within the window size, R (Figure S3A). R is the number of row and column elements additional to the diagonal elements of a cumulative cost matrix $\mathbf{D}^{\mathbf{N}\times\mathbf{M}}$(3). The Itakura Parallelogram sets the slope of the boundary that makes up the windowing constraints (see Figure S3B)(8). We selected the Sakoe-Chiba windowing constraint because it imposes a constant restriction on the selection of warping path, unlike the Itakura Parallelogram (see Figure S3). It also allows users to pre-specify a window size (see Figure S4), which we specified at 28 days for this study. Sakoe-Chiba windowing constraints with size of 28 days prevent the DTW algorithm from matching time points that are more than 28 days apart. To illustrate the differences in DTW output when windowing constraints are altered, we also performed a DTW analysis without windowing constraints, and the results are attached in Supplementary 5.

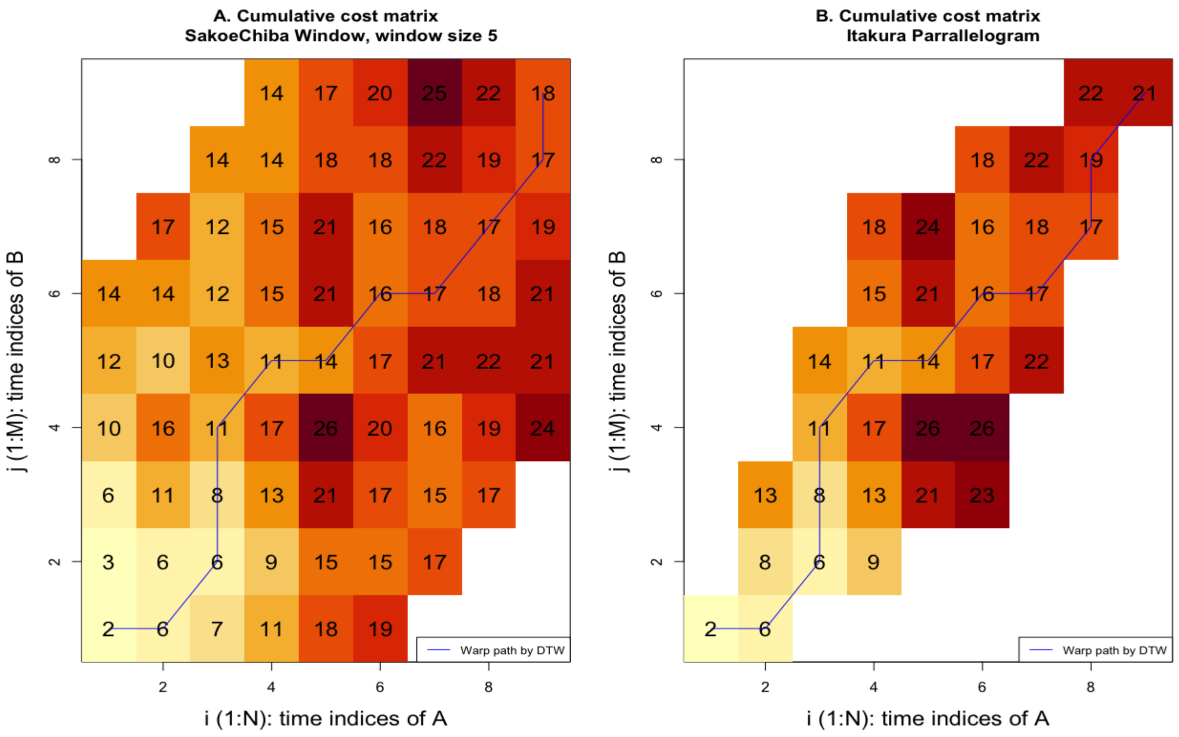


**Figure S3: Windowing constraints**

Two common windowing constraints: (Left) Sakoe-Chiba window with window size (R) of 5 imposed on the cumulative cost matrix of the DTW analysis of A and B. In both directions of i and j, R is the five elements in addition to the diagonal element when i = j. (Right) Itakura Parallelogram imposed on the cumulative cost matrix from the DTW analysis of A and B.

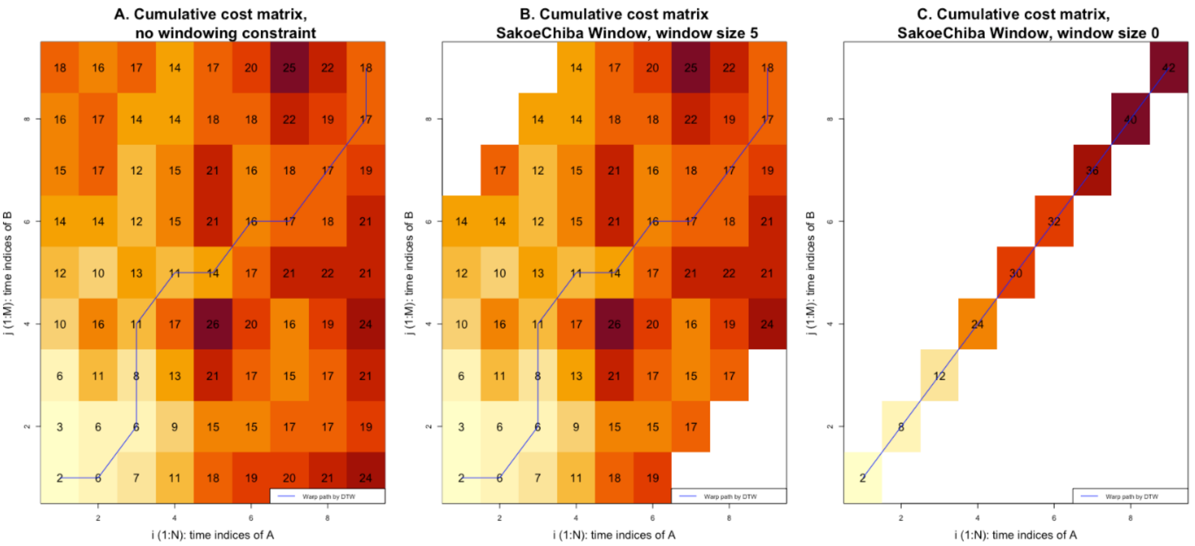


**Figure S4: Window size**

All three figures have the DTW step patterns pre-specified to symmetric 2. The blue lines represent the optimal warping path computed by DTW. The value in the top right cell (N, M) shows the cumulative cost of the whole path. **Figure S4A** (Left): Cumulative cost matrix with no windowing constraints. **Figure S4B** (Middle): Cumulative cost matrix with Sakoe-Chiba window with window size of 5. **Figure S4C** (Right): Cumulative cost matrix with Sakoe-Chiba window with a window size of 0. Figure S4C has a window size of 0, essentially forcing the warping path to be on the diagonal elements, i.e., only elements of the same time indices from A and B are matched, and their differences in A and B values were calculated. Note that the optimal warping path and final cost differ between Figure S4A and S4C but not between Figure S4A and S4B. This difference demonstrates how window size affects the determination of the optimal warping path and the computed distance between time series.

#### Example 1: DTW analysis of A and B using cumulative cost matrix $\mathbf{D}^{\mathbf{N}\times\mathbf{M}}$ and step pattern symmetric2(3,5)

Using the time series A and B as an example, we outlined how the DTW algorithm:

1. Constructs a cumulative cost matrix, $\mathbf{D}^{\mathbf{N}\times\mathbf{M}}$, (see Figure S5) under two constraints: step patterns and windowing constraints
2. Computes the DTW distance
3. Identifies the optimal warping path

The example has a pre-specified symmetric2 for step pattern with no windowing constraints.

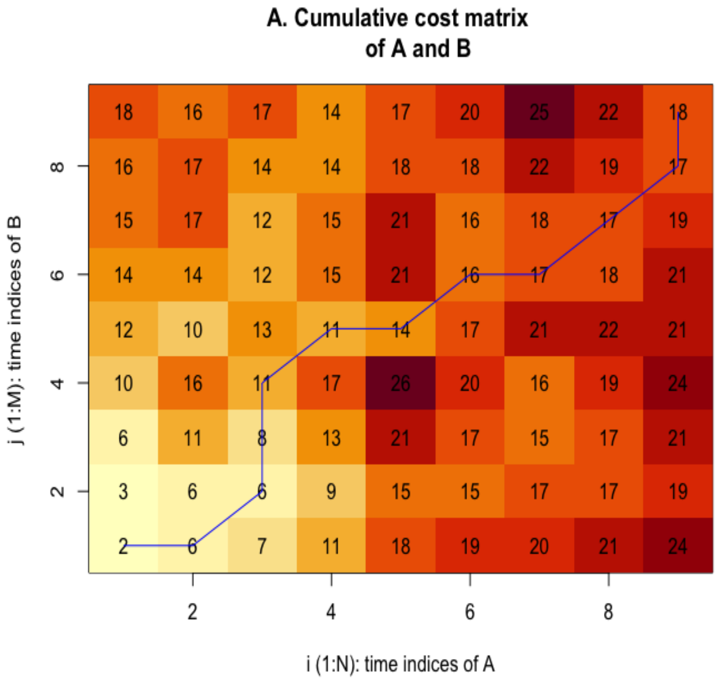


**Figure S5:** The cumulative cost matrix of A and B, $\mathbf{D}^{N \times M}$. Blue line represents the optimal warping path.

<u>Step 1 to 3 demonstrate how $\mathbf{D}^{N \times M}$ (Figure S5) is built:</u>

1. For each element in the first row of the matrix $\mathbf{D}^{N \times M}$, the value is calculated by: $D(i,1) = \sum_{k=1}^{i} d(a_k, b_1), i \in [1, N]$. Where,
   a. $D(i,1)$ is the cumulative cost of the element in the first column.
   b. $d(a_k, b_1)$ represents the differences in value between first value of B and the $i^{th}$ value of A.
   c. For example, $D(4,1)$ = 11, this is calculated from:
      i. Finding the absolute difference between $a_4$ and $b_1$, $d(a_4, b_1)$: $9 - 5 = 4$.
      ii. Adding $d(a_4, b_1)$ to the value in cell $D(3,1)$, 7, the cumulative cost from the 1st to 3rd cell in the first row.
      iii. The final cumulative cost for cell (4,1) is 11.
2. For each element in the first column of the matrix $\mathbf{D}^{N \times M}$, the value is calculated by: $D(1,j) = \sum_{k=1}^{j} d(a_1, b_k), j \in [1, M]$. Where,
   a. $D(1,j)$ is the cumulative cost of the $j^{th}$ elements in the first row.
   b. $d(a_1, b_k)$ represents the differences in value between first value of A and the $j^{th}$ value of B.
   c. Thus, for the element with coordinate (1,4), the value is derived by:
      i. Finding the absolute difference between $b_4$ and $a_1$, $d(a_1, b_4)$: $|3 - 7| = 4$.
      ii. Adding $d(a_1, b_4)$ to $D(1,3)$, 6, the cumulative cost from the 1st to 3rd element in the first column.
      iii. Thus, the final cumulative cost for element (1,4) is 10.

3. For all the other elements $D(i, j)$, the values are computed as below as per the step pattern, symmetric2:

$$D(i,j) = \min \begin{cases} D(i-1, j-1) + 2 \times d(a_i, b_j); \\ D(i-1, j) + d(a_i, b_j); \\ D(i, j-1) + d(a_i, b_j) \end{cases}$$

Thus, for the element with coordinate (5, 5), the value $D(5,5) = 14$, is derived by:

a. Finding the absolute difference between the $a_5$and $b_5 = |12 - 9| = 3$.

b. Calculating the cost for these three scenarios and choosing the value with the lowest value as $D(5,5)$.

   i. $D(i-1, j-1) + 2 \times d(a_5, b_5) = D(4,4) + 2 \times 3 = 17 + 6 = 23$

   ii. $D(i-1, j) + d(a_5, b_5) = D(4,5) + 3 = 11 + 3 = 14$

   iii. $D(i, j-1) + d(a_5, b_5) = D(5,4) + 3 = 26 + 3 = 29$

c. $D(i-1, j) + d(a_5, b_5)$ has the lowest value, thus $D(5,5)$ = 14

Step 4 demonstrates how DTW distance is computed:

1. We can use $\mathbf{D}^{\mathbf{N \times M}}$ to calculate the DTW distance between A and B using the formula below, under the restrains of symmetric2 step pattern:

$$DTW(A,B) = \min \begin{cases} D(N-1, M-1) + 2 \times c(a_N, b_M) \\ D(N-1, M) + c(a_N, b_M) \\ D(N, M-1) + c(a_N, b_M) \end{cases}$$

$$= \min \begin{cases} D(8,8) + 2 \times d(a_9, b_9) \\ D(8,9) + d(a_9, b_9) \\ D(9,8) + d(a_9, b_9) \end{cases} = \min \begin{cases} 19 + 2 \times 1 \\ 22 + 1 \\ 17 + 1 \end{cases} = 18$$

Step 5 demonstrates how the optimal warping path is determined:

1. The optimal warping path is determined by backtracking from $p_{end} = (N, M)$ to $p_{start} = (1,1)$. The algorithm determines the direction of this path by:

   a. Backtracking from $p_{end} = (N, M)$.

   b. Selecting the element with the lowest cumulative cost. These elements can be:

      i. Horizontally left of $(N, M)$, i.e. $(N-1, M)$, or

      ii. Diagonally across $(N, M)$, i.e. $(N-1, M-1)$, or

      iii. Vertically below $(N, M)$, i.e. $(N, M-1)$.

   c. Repeating step 5b for each newly chosen element until the path reaches $p_{start} = (1,1)$.

### Supplementary 2 (Methods – relative time lag)

The DTW optimal warping path between two time series also describes their lead-lag time relationship. To quantify this relationship, we calculated two parameters from the output of DTW algorithm: average time lag (ATL) and relative time lag (RTL).

#### Average time lag (ATL) and the lead-lag time relationship between two time series

ATL is a parameter that quantifies the mean lead-lag time relationship between two time series. We derived ATL using the optimal warping path identified via DTW. ATL for any given period can be extracted from the warping path. We illustrate this further using time series X and Y in Example 2. X and Y are two time series with the same values and shapes, except X is five days ahead of Y (see Figure S6). Figure S7 shows their optimal warping path. The ATL of X and Y is calculated by obtaining the difference between the optimal warping path ($p^*$) and the diagonal line (q) where the time indices are equal (see Example 2)

##### Example 2: Average time lag (ATL) between X and Y for a defined period, $\boldsymbol{\alpha}$

Step 1: Finding $\delta_l$ and the coordinate $(q_i, q_j)$ (see Figure S7C):

Where $\delta_l$ (red line) is the perpendicular distance between the points on $p^*$ and q:

1. Optimal warping path, $p^* = (p_1, p_2, \ldots, p_K)$ with ${p^*}_l = (p_i, p_j) \in [1:N] \times [1:M]$ for $l \in [1:K]$, and
2. $q = (q_1, q_2, \ldots, q_K)$ with $q_l = (q_i, q_j) \in [1:N] \times [1:M]$ for $l \in [1:K]$, q is where $\delta_l$ join the diagonal line (green)

$$\delta_l = \sqrt{(p_i - q_i)^2 + (p_j - q_j)^2}\text{, and because } m = n\text{, thus } q_j = q_i$$

$$\delta_l = \sqrt{(p_i - q_i)^2 + (p_j - q_i)^2}, \qquad {\delta_l}^2 = (p_i - q_i)^2 + (p_j - q_i)^2,$$

$$\frac{\partial {\delta_l}^2}{\partial q_i} = -2(p_i - q_i) - 2(p_j - q_i)$$

$$\frac{\partial {\delta_l}^2}{\partial q_i} = 0\text{, given } \delta \text{ is the minimum distance between warping path, p, and q}$$

Thus, $-2(p_i - q_i) - 2(p_j - q_i) = 0$; and $q_i = \frac{p_i + p_j}{2}$, and $q_j = \frac{p_i + p_j}{2}$

Now that we know $q_i = \frac{p_i + p_j}{2}$, and $q_j = \frac{p_i + p_j}{2}$

$$\delta_l = \sqrt{\left(p_i - \frac{p_i + p_j}{2}\right)^2 + \left(p_j - \frac{p_i + p_j}{2}\right)^2}, \text{ and } \delta_l = \sqrt{\left(\frac{p_i - p_j}{2}\right)^2 + \left(\frac{p_j - p_i}{2}\right)^2}$$

Step 2: Finding the average of $\delta_l$, which is the ATL between X and Y:

$$\mathrm{ATL(X, Y)} = \frac{\sum_{l=1}^{K} \delta_l}{\mathrm{K}},$$

Where:

1. $\sum_{l=1}^{K} \delta_l$ is the sum of all perpendicular distance $\delta_l$.
2. K is the total number of $\delta_l$.

Step 3: Obtaining the lead-lag time graph (Figure S7D):

The lead-lag time graph is analogous to rotating the warping path 45 degrees to the right.

The values for y and x-axis for the lead-lag time graph are:

1. y-axis comprises the values of all the perpendicular distance, $\delta_l$.
2. x-axis comprises the values along the diagonal $(q_l)$ which $\delta_l$ map to. These values are the $q_i$ or $q_j$ coordinate of the diagonal from the warping path graph. This is analogous to the average time at each step of realignment between X and Y.

Step 4: Extracting a period-specific ATL, $\mathrm{ATL(X, Y)}_t$, using the lead-lag time graph:

We can extract the ATL for a specific period of analysis based on the x-axis (t) in the lead-lag time graph in Figure S7D. For example, for a period of interest, α, between time a and b, where $a \le t \le b$:

$$\mathrm{ATL(X, Y)}_\alpha = \frac{\sum_a^b \delta_\alpha}{\mathrm{K}_\alpha},$$

Where:

1. $\sum_a^b \delta_\alpha$ is the sum of all δ in the period of interest α, $a \le t \le b$.
2. $\mathrm{K}_\alpha$ is the total number of δ in the period of interest α, $a \le t \le b$.

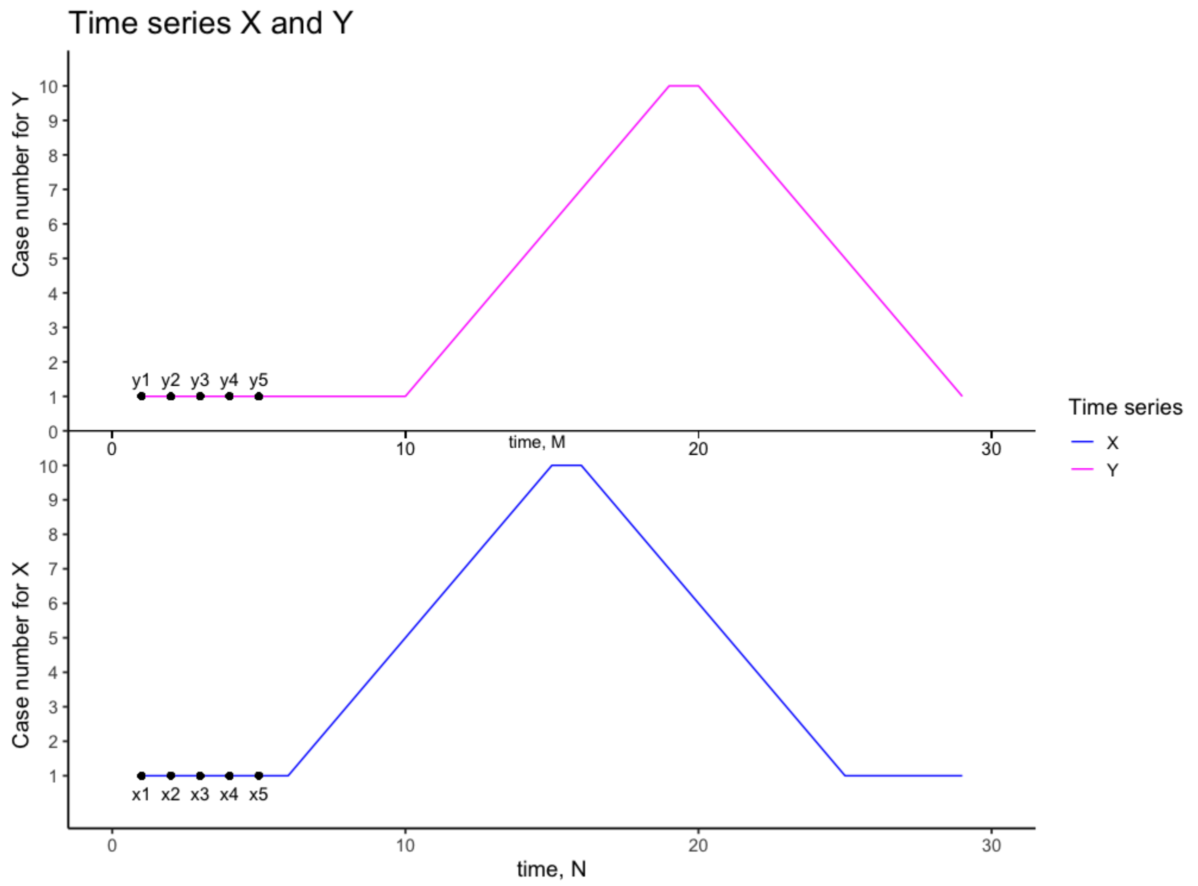


**Figure S6: Time series X and Y**

Time series X (blue) and Y (pink) are plotted above one another. X (blue) and Y (pink) are two time series with the same values and shapes, except X is five days ahead of Y.

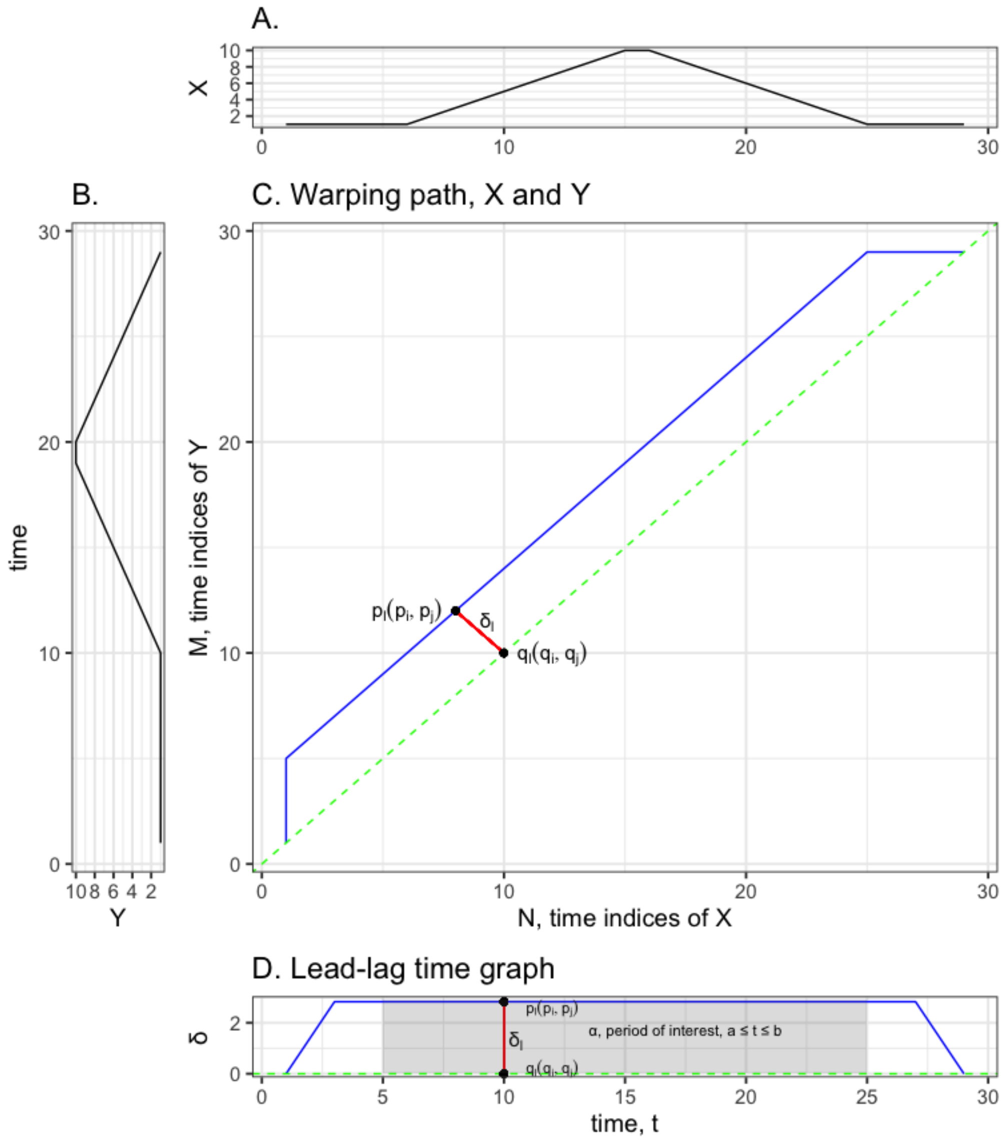


**Figure S7: Optimal warping path and lead-lag time graph of X and Y**

DTW analysis and optimal warping path of X and Y: **Figure S7A** and **Figure S7B** are the time series X and Y. **Figure S7C** shows the optimal warping path, $p^*$, in blue, and the diagonal line, $q$, in green. $\delta_l$ is the perpendicular distance between a point in the optimal warping path, $p_l = (p_i, p_j)$, and the diagonal line $q$. Where $\delta$ meets $q$ is a sequence of points with coordinates of $(q_i, q_j)$. The number of $\delta_l$ depends on the number of points on the optimal warping path, $p_l$. We use the formula in Example 2 to calculate the $\delta$ and $(q_i, q_j)$ for each point of the warping path $(p_i, p_j)$. Using the calculated $\delta$ and $(q_i, q_j)$, we calculated an average value for $\delta_l$, the ATL, for X and Y. **Figure S7D** is the lead-lag time graph for X and Y, and the grey-shaded area demonstrates the period of interest, $\alpha$. ATL specific to $\alpha$, can be calculated by limiting the averaging of $\delta_l$ to this period only.

#### Relative time lag (RTL) and the lead-lag time relationship between one time series and several (more than one) time series

When performing lead-lag time analysis comparing a large number of time series, it is not feasible to present every pairwise ATL. Thus, we used another indicator, relative time lag (RTL), to quantify the lead-lag time relationship between a time series and all other time series it compares to. Time series with a positive RTL lead other time series, while time series with a negative RTL lag other time series. The degree of time-lead, or time-lag, increases as RTL moves further away from zero. RTL differs from ATL in that RTL compares one time series to more than one time series. In contrast, ATL compares one time series to another time series. We can also calculate the RTL for each time series across different analysis periods. We demonstrate this in Example 3.

##### Example 3: Relative time lag (RTL) for time series $S_1$ in a set of time series, $S$

Let a set of time series be $S = \{S_1, S_2, S_3, \dots, S_a\}$, where, $a \in 1: W$, $W$ is the total number of time series in set S, and a is the $a^{th}$ time series in set S

Let $ATL_{S_1,\alpha,z}$ be the list of average time lag (ATL) values for a time series, $S_1$, across a period, $\alpha$, when compared individually to each of the remaining time series in $S, z$:
$ATL_{S_1,\alpha,z} = \{ATL(S_1, S_2)_\alpha,\ ATL(S_1, S_3)_\alpha \dots\ ATL(S_1, S_z)_\alpha\}$, where $z\ \in a\ \in 1: W$, but $z \neq 1$

The RTL for a time series $S_1$ across duration $\alpha$, when compared to all the remining time series is:

$$RTL_{S_1,\alpha} = \frac{\sum_{z=2}^{W} ATL_{S_1,\alpha,z}}{W-1}, \text{where}$$

1. $\sum_{z=2}^{W} ATL_{S_1,\alpha,z}$ is the sum of all elements in $ATL_{S_1,\alpha,z}$
2. $W - 1$ is the number of time series in set $S$ $S_1$ is comparing to.

### Supplementary 3 (Methods – lead-lag time analysis)

#### Lead-lag time analysis of COVID-19 time series from 380 LTLAs in the UK

We performed DTW analysis to investigate the lead-lag time relationship amongst 380 time series in the UK for four periods of interest (see Figure 1 for an example of pairwise DTW analysis of time series from two LTLAs). These periods are defined as:

1. 29 November 2020 to 21 April 2022: This period was obtained after excluding the first and last 28 days of the warping path computed by pairwise DTW analysis of time series. This exclusion accounts for DTW's methodological limitation of accurately matching the pairwise time series at the start and the end of the time series.
2. 29 November 2020 to 21 March 2021: This period was when the Alpha variant was emerging and became dominant. The Alpha variant was first detected in September 2020. We selected 29 November 2020 as that was the earliest data available for analysis after accounting for missing data in October 2020 and excluding the first 28 days.
3. 28 April 2021 to 18 August 2021: This period was when the Delta variant was emerging and became dominant.
4. 29 November 2021 to 21 March 2022: This period was when the Omicron BA.1 variant was emerging and became dominant.

The start dates for the Delta and Omicron BA.1 variants were selected at the point when more than 1% of the sequenced COVID-19 cases belonged to the respective variant(9). For periods 2 to 4, the end dates were 16 weeks after the start date, assuming it takes 16 weeks for each variant to spread through the UK. Using the method in Example 2 and 3, we obtained four period-specific RTLs for each time series. We then visualised the period-specific RTLs on a map of LTLAs.

#### Lead-lag time analysis of COVID-19 time series within each nation

Using the same definition for the periods of interest as in the UK lead-lag time analysis, we also analysed the lead-lag time relationship of the time series from each of the four nations separately. Within each of these four nation-specific lead-lag time analyses, the DTW analysis was limited to: (1) 315 time series from LTLAs in England; (2) 22 time series from LTLAs in Wales; (3) 11 time series from LTLAs in Northern Ireland; and (4) 32 time series from LTLAs in Scotland respectively. We obtained four period-specific RTLs for each of the LTLA's time series. We then visualised the period-specific RTLs on LTLA maps of each nation.

### Supplementary 4: Lead-lag time analysis with windowing constraints for Scotland, Wales and Northern Ireland

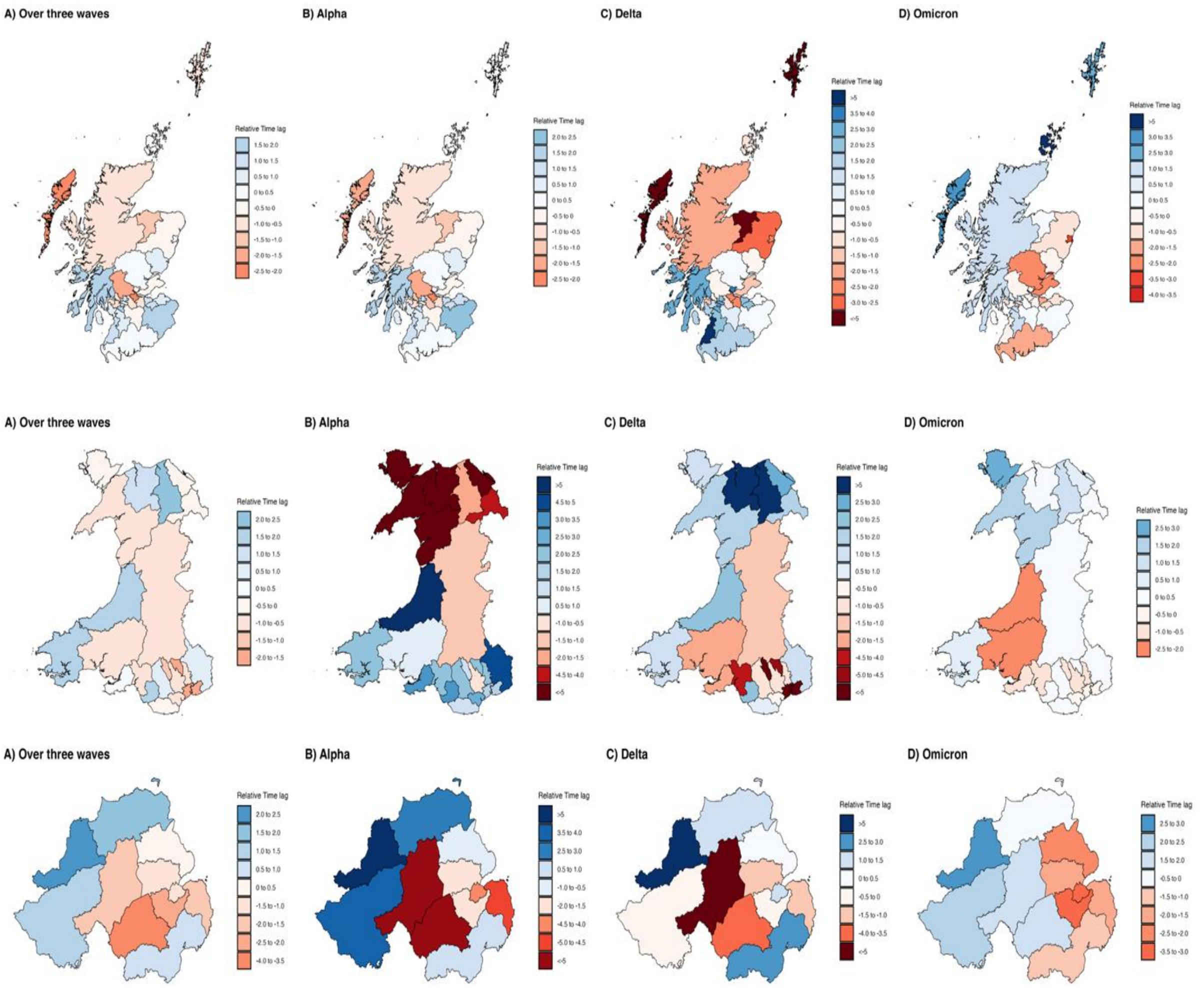


**Figure S8: Lead-lag time analysis for Scotland, Wales and Northern Ireland.**

Scotland (top), Wales (centre) and Northern Ireland (bottom) analysed across four analytic periods.

### Supplementary 5: Hierarchical clustering analysis and lead-lag time analysis without windowing constraints

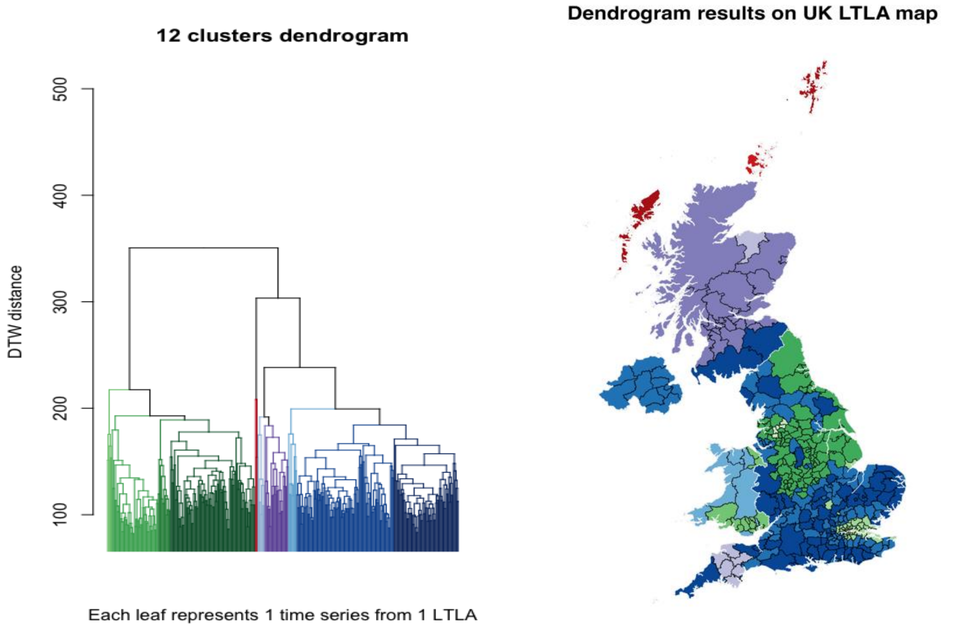


**Figure S9: Hierarchical clustering analysis without windowing constraints**

The output of hierarchical clustering analysis divided into 12 clusters

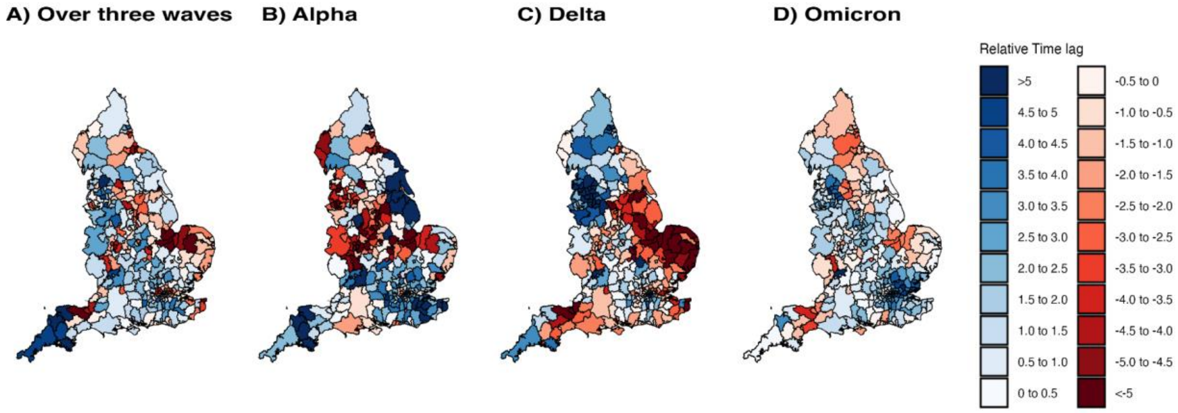


**Figure S10: Lead-lag time analysis in England without windowing constraints across four analytic periods**

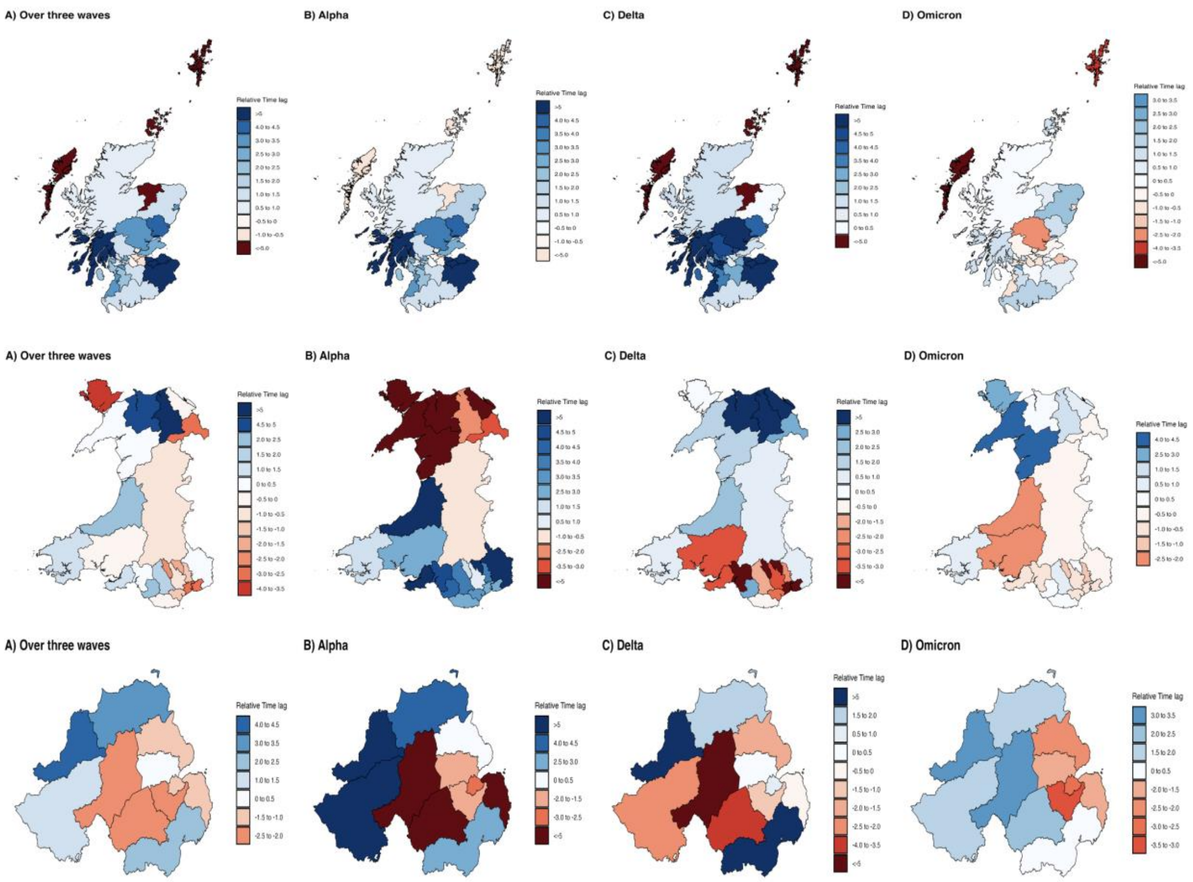


**Figure S11: Lead-lag time analysis in Scotland, Wales, and Northern Ireland without windowing constraints across four analytic periods.**
